# The Study Protocol for GENESIS: GENEral population normS—An International Survey

**DOI:** 10.64898/2026.01.27.26344920

**Authors:** Sarah Dewilde, Nafthali H. Tollenaar, Glenn Phillips, Sandra Paci, Clémence Arvin-Berod

## Abstract

**Background:** Chronic autoimmune diseases such as Chronic Inflammatory Demyelinating Polyneuropathy (CIDP), Multifocal Motor Neuropathy (MMN), and Thyroid Eye Disease (TED) impose a considerable burden on affected individuals. Patient-reported outcome measures (PROMs)—both disease-specific and generic—are widely used to assess functioning, quality of life, and treatment effects in these populations. However, most PROMs currently lack reference values derived from the general population, limiting the interpretability of patient scores.

**Objective:** The GENESIS (GENEral population normS—An International Survey) study aims to establish general population norms for a range of PROMs used in CIDP, MMN, and TED across six countries: Germany, Italy, Japan, Spain, the United Kingdom, and the United States. These norms will improve patient score interpretation and help quantify unmet needs in patients with these rare autoimmune diseases.

**Methods:** GENESIS is an observational, cross-sectional, online survey of the adult general population (N=21,000). Participants will be recruited to be representative by age, gender, region, and education. The survey includes validated instruments such as the EQ-5D-5L, I-RODS, MMN-RODS, CAP-PRI, GO-QoL, BPI-SF, RT-FSS, FACIT-Fatigue, HADS, and WPAI, along with items on demographics, caregiver need, and healthcare utilization. To reduce respondent burden, participants will be randomized into two groups, each completing a subset of the full questionnaire. A subset of respondents (n=2,333) will be re-surveyed after two months to support psychometric validation. Data will be analyzed descriptively to generate normative values for each PROM by country and in aggregate.

**Results and Dissemination:** Data collection is scheduled to begin in August 2025, with results expected by Q4 2025. Findings will be disseminated via peer-reviewed publications and conference presentations.

**Conclusion:** GENESIS will provide foundational normative data across six countries for PROMs commonly used in rare autoimmune diseases. These data will support more meaningful interpretation of PROM scores in both clinical practice and research settings.

## 1 Background

Patients with chronic autoimmune diseases, such as Chronic Inflammatory Demyelinating Polyneuropathy (**CIDP**), Multifocal Motor Neuropathy (**MMN**), and Thyroid Eye Disease (**TED**), experience a substantial burden due to the progressive and debilitating nature of their symptoms^1–4^. However, due to the rarity of these conditions, this burden is often less recognized than that of more common diseases, and treatment options remain limited. To assess and monitor disease progression, patients frequently complete validated generic and disease-specific patient-reported outcome measures (PROMs) during both routine and acute healthcare visits. These PROMs also serve as primary endpoints in clinical trials—providing insights into patient functioning and treatment efficacy.

In CIDP, commonly used PROMs include the Inflammatory Rasch-Built Overall Disability Scale (**I-RODS**) and the Chronic Acquired Polyneuropathy Patient-Reported Index (CAP-PRI). In MMN, the **MMN-RODS** is frequently administered, while TED patients often complete the Graves’ Ophthalmopathy Quality of Life (**GO-QOL**). Additionally, instruments like the **EQ-5D-5L**, Rasch-Transformed-Fatigue Severity Scale (**RT-FSS**), Brief Pain Inventory – short form (**BPI-SF**), Hospital Anxiety and Depression Scale (**HADS**), and the Work Productivity and Activity Impairment Questionnaire (**WPAI**) are also relevant in these patient populations.

Most disease-specific PROMs lack reference values derived from the general population. **Establishing general population norms for these outcomes provides an important baseline for interpreting scores within disease populations, enabling a clearer assessment of disease burden and facilitating comparison with broader health benchmarks**.

## 2 Study objectives

This survey aims to generate general population norms in 6 countries (Germany, Italy, Japan, Spain, United Kingdom [UK], and the United States [US]) for PROMs that are measured in CIDP, MMN, and TED, to improve interpretation of scores and help to quantify unmet needs of patients.

## 3 Study design

This is a digital, observational, multi-country survey among the general population. Data will be collected via an online self-administered survey. All participants will complete the survey once during the initial and main data collection (wave 1). Approximately 2-3 months after wave 1, a small subset of participants will be invited a second time to complete a shorter version of the survey (wave 2). The data of the second wave will be used to measure changes over time, and test psychometric properties of several instruments.

## 4 Study population & Sample size TOTAL 21,000 2,333

### 4.1 Wave 1

In the first wave, data will be collected among 21,000 adult members of the general population in 6 countries: 6,000 in the US, and 3,000 in Germany, Italy, Japan, Spain, and the UK (Table 1). The sample size is not based on a formal calculation but driven by the aim to be representative of the general population, and by feasibility. A large sample size will be collected in US to account for its large and diverse population and unsure representative estimates. The only inclusion criteria for participants is the minimum age of 18 years old. It is estimated that the period needed to obtain results of 21,000 individuals will be three to six months.

**Table 1.**
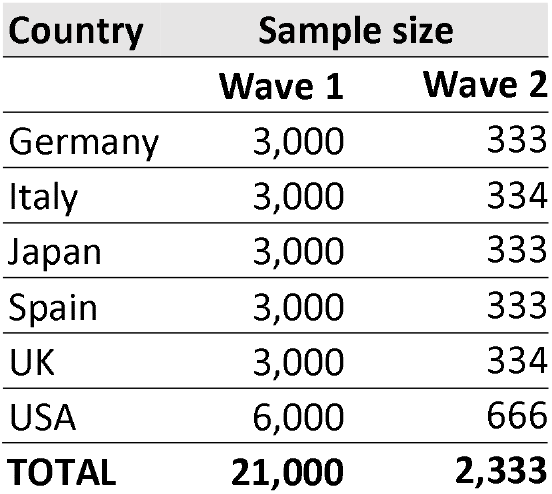
Sample size per country.

### 4.2 Wave 2

In the second wave, data will be collected among 2,333 members of the general population: 667 in the United States, and 333 in Germany, Italy, Japan, Spain, and the UK (Table 1). The only inclusion criterion for participants is the minimum age of 18 years old. Wave 1 respondents will be stratified by health severity to ensure adequate representation across symptom levels. After analyzing potential anchor variables and their correlations with the PROMs, the rescaled (0-100) EQ-5D-5L Level Sum Score (LSS) was identified as the most suitable stratification measure, using the following categories: no problems (LSS=0), mild (LSS=1–19), moderate (LSS=20–39), severe (LSS=40–59), and extreme (LSS=60–100).

## 5 Data collection

The full survey will be programmed in Qualtrics (version April 2025). All questions will be optimized to be displayed on both desktop and mobile devices. To avoid having large quantities of missing data, respondents are required to answer all questions. Because of this, there will be no missing data, unless participants decide not to finish the survey.

The survey will start by collecting online informed consent (**Supplementary file 1**), followed by a Captcha test. First survey items will be demographic questions and a measure of health-related quality of life (EQ-5D-5L). Next, respondents will complete a subset of clinical outcome measures (e.g. I-RODS or the GO-QOL), and PROMs on pain, fatigue, and mental health. After that, some questions will be asked about people’s need for a caregiver and their use of medical resources in the past month. The study will end with asking about work productivity. Respondents do not have to complete all instruments, but are randomized into two groups, each covering a subset of questions. Each section of the questionnaire is detailed below.

### 5.1 Demographics

- **Age** in years
- **Sex** (i.e., male, female or other)
- **Marital status** (i.e., married/partnered, single, or widowed)
- **Current living situation** (i.e., I live in my own home / apartment; with my partner and/or children; I live in my family home / apartment; with my parents, I live in my own home / apartment, alone; I live in the home / apartment of a family member that are not parents; I live in a co-housing / with roommates; I live in a nursing home; I live in a long-term care rehabilitation facility)
- **Level of education** (i.e., primary school, secondary school, and higher education)
- **Current work situation** (i.e., Employed (full-time); Employed (part-time); Independent or self-employed; Student; Unemployed; Retired; On long-term disability leave)
- **Weight** (in lbs/kg), height (in ft/cm)
- **Use of mobility aids** (i.e., a cane, a frame, crutches, a walker or a wheelchair)
- **Presence of co-morbidities** (i.e., Cardiovascular disease (e.g. coronary heart disease, stroke); Respiratory disease (e.g. asthma, COPD, emphysema, bronchitis); Stomach or peptic ulcer; Liver disease; Kidney disease; Skin disease (e.g. psoriasis, vitiligo); Inflammatory bowel syndrome/disease (e.g. Crohn’s disease, ulcerative colitis); Cancer (e.g. breast cancer, prostate cancer); Arthritis; Osteoporosis; Other musculoskeletal disease (e.g. bursitis, sarcopenia, osteopenia); Multiple sclerosis; Other autoimmune disease (e.g., lupus, myasthenia gravis, Guillain-Barré syndrome, CIDP, Graves’ disease, Addison disease, pernicious anemia); Thyroid disorder/disease; Diabetes; Obesity; Hearing loss; Visual impairment (e.g. cataract, glaucoma); Sleep disorder; Migraines; Epilepsy; Long COVID; Alzheimer’s disease or dementia; Anxiety; Depression; Other mental health disorders; Eating disorders (e.g., bulimia, anorexia nervosa); None of the above)

### 5.2 Validated instruments: Health-related Quality of life and functioning

1. EQ-5D-5L: The EQ-5D-5L, a widely used instruments to value health, includes five questions covering mobility, self-care, usual activities, pain/discomfort and anxiety/depression. At the end there is a visual analogue scale (VAS) from 0 (worst imaginable health) to 100 (best imaginable health)^5^.
2. EQ-5D Bolt-ons: Five additional dimensions (on Tiredness, Energy, Sleep, Fatigue, and Exhaustion) will be added to the standard EQ-5D-5L, plus a ranking exercise.
3. EQ-5D modified dimensions: The composite dimensions of the EQ-5D-5L (Pain/Discomfort, Anxiety/Depression) and EQ-5D-Y-5L^6^ (Worried/Sad/Unhappy) will be included as composite and split-up items. A mirrored dimension of ‘Unhappy’ (Happy’) is also added.

### 5.3 Validated instruments: Clinical Outcomes

4. I-RODS: The Inflammatory Rasch-Built Overall Disability Scale specifically designed for assessing activity limitations in individuals with CIDP^7^.
5. MMN-RODS: The Rasch-Built Overall Disability Scale for Multifocal Motor Neuropathy is similar to the I-RODS but tailored to the specific functional limitations experienced by MMN patients, providing a score that reflects their daily activity challenges^8^.
6. CAP-PRI: The Chronic Acquired Polyneuropathy Patient-Reported Index is a patient-reported outcome measure developed to assess the functional impact of symptoms in patients with CIDP, providing insight into the physical and emotional burden of the disease^9^.
7. GO-QoL: The Graves’ Ophthalmopathy Quality of Life is specifically developed for TED patients. It assesses the impact of eye symptoms on visual functioning and psychosocial aspects, including self-confidence and social interactions^10^.

### 5.4 Validated instruments: Other PROMs

8. BPI-SF: The Brief Pain Inventory short form includes questions on pain location, intensity, relief, and how pain interferes with activities like mood, walking, and sleep^11^.
9. RT-FSS: The Rasch-Transformed Fatigue Severity Scale, provides a robust measure of fatigue severity with questions focused on the impact of fatigue on daily activities^12^.
10. FACIT-Fatigue: The Functional Assessment of Chronic Illness Therapy – Fatigue is a 13-item measure that assesses self-reported fatigue and its impact upon daily activities and function^13^.
11. HADS: The Hospital Anxiety and depression scale is a simple and reliable tool to assess anxiety and depression, including seven items reflecting depression and seven reflecting anxiety^14^.

### 5.5 Work productivity

12. WPAI: The Work Productivity and Activity Impairment Questionnaire is designed to measure the impact of health problems on work productivity and daily activities .

### 5.6 Caregiver need and medical resource utilization

- Need for a caregiver
- Use of medical services used during the past month (hospital, general practitioner, specialist)

## 6 Adaptation of PROMs to the general population

All clinical outcomes included in this study (I-RODS, MMN-RODS, CAP-PRI, GO-QOL) were specifically developed and validated for use in patients with CIDP, MMN or TED. Where necessary, minor wording adjustments were made to ensure the questions are appropriate and understandable for the general population. Adjustments were made carefully, conservatively and consistently, to stay as close as possible to the original instruments. In particular, disease-specific references (e.g., “your polyneuropathy”) were replaced with the phrase “an ongoing health problem”. This phrasing was selected for its neutrality, broad applicability, and ability to discourage respondents from referring to short-term issues (e.g. a broken leg or headache). Examples of how the wording was adjusted are provided in **Appendix 1**.

## 7 Survey flow

To reduce the total number of questions to be answered per respondent in wave 1, participants will be split into two groups and randomized between one of two sets (**Figure 1**). Group 1 will complete the I-RODS and CAP-PRI, while group 2 will complete the MMN-RODS, and GO-QOL questionnaire. Additionally, respondents will complete either one of two sets of additional questions to the EQ-5D, plus either the FACIT-fatigue or the RT-FSS. This reduces the total number of questions from 176 for all participants, to 121 in group 1 and 118 in group 2. Based on our previous experience with this sort of surveys, we estimate that completing the survey will take 12 minutes on average. (min. 8, max. 18 minutes).

**Figure 1.**
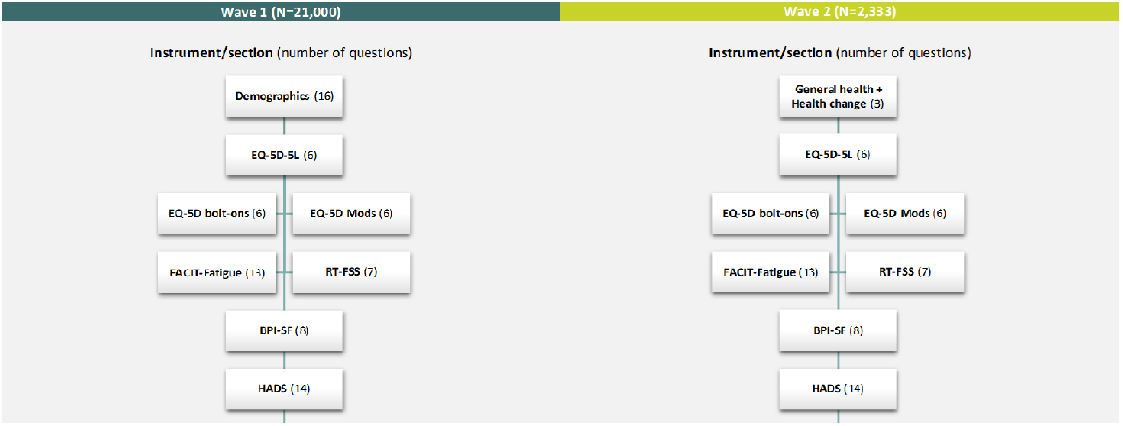
Survey Flow chart of waves 1 and 2 of the GENESIS study

In wave 2 (**Section 4.2**), all participants will complete several questions about their overall health, change in health, and complete all the same PROMS that were included in wave 1, to allow for comparison. To ensure that participants in Wave 2 completed the same questionnaire blocks as in Wave 1, each respondent’s original block assignments were recorded and transferred to Qualtrics via query parameters as embedded data fields. These fields guided the survey logic to route participants to their corresponding blocks, maintaining consistency and comparability across waves.

## 8 Analysis

The study is purely descriptive. There is no formal hypothesis that we aim to test. Descriptive statistical methods will be used to analyze and describe the data set. Pooled results for all the countries will be presented as well as results per country. Population standards for each of the PROMs, individual items and total scores will be estimated. The statistical analysis software programs SAS and R will be used for analysis.

In this study, an experimental arm was added specifically to investigate the psychometric properties of the EQ-5D-5L composite dimensions (comparing combined and single items), and five vitality related EQ-5D-5L bolt-ons: tiredness, energy, fatigue, exhaustion, and sleep. A detailed description of rationale and methodology is added in **Appendix 2**.

## 9 Survey conduct and procedures

### 9.1 Recruitment

The sample will be drawn from a representative panel of administered by Bilendi and trusted panel partners for Japan and the US. These panels include 150,000 willing survey participants with pre-specified profiles based on over 300 criteria. The panel of respondents is constantly being updated, and respondents are recruited in the panel from various sources (TV, radio, newspapers), and different titles per medium to ensure a suitable representation of each country’s population. Only panel members can participate in the survey; no river sampling, routing or real-time web sampling will be used.

Bilendi employs proportionally stratified random sampling (interlaced quotas) to ensure representativeness on age, gender, region, and education. Propensity weighting is used post hoc to adjust the sample distribution, particularly to correct for potential underrepresentation of groups such as older women or those with lower education levels.

Potential participants are invited to participate through personalized e-mail invitations and reminders containing a unique link to the questionnaire. A micro-sending strategy is employed to spread invitations over several days and times (including weekends), minimizing bias from overly online or reactive participants.

### 9.2 Incentive

Participants will receive points from Bilendi, the market research company administrating the online panel, which can be exchanged for gifts. The length of the survey will earn the participants approximately 150 points, which is less than 1 euro. This modest reward ensures that the participant pool is composed of individuals motivated by interest and engagement, not incentives alone.

### 9.3 Soft launch

The initial phase of data collection will follow a stepwise approach, effectively serving a pilot function. During this phase, response patterns will be monitored, data quality will be checked, and issues related to survey design or translations will be identified and resolved. Data collected during this phase may still be included in the main analysis, provided no substantial changes to the survey are required.

### 9.4 Monitoring data quality

During the initial inclusion of new participants to the online panel, checks include GeoIP, browser/device fingerprinting, duplicate detection, double opt-in confirmation, and Captcha validation. Disposable email domains are screened and flagged automatically.

During survey completion, additional checks include:

- Completion time monitoring: speeders are flagged if their completion time is less than 5 minutes.
- A control (trap) question to identify inattentive respondents (‘Please select B as your answer’).
- Duplicate cookie and device checks are performed.

Online panel participant’s behavior is tracked across Bilendi surveys through a dynamic quality scoring system. Panel members demonstrating poor response quality (e.g., straight-lining, inconsistent answers) are banned quarterly.

### 9.5 Data storage, security, and ownership

Data will be collected though the Qualtrics platform and securely stored on servers managed by CHEOPS, in accordance with applicable data protection regulations (GDPR and NIS2). Only authorized research will have access to raw data. Services in Health Economics (SHE) is the sole owner of all collected data in this study.

### 9.6 Timelines

It is planned to start data collection in August 2025 and end it until all data have been collected, 3 to 6 months later. In Q4 2025, initial results will be published.

### 9.7 Publication and data sharing policy

Survey results will be published in peer-reviewed articles, aiming towards high impact journals.

## Data Availability

All data produced in the present study are available upon reasonable request to the authors.

## 9.9 Ethical considerations

Individuals can only complete the online survey after agreement of the online informed consent form (**ICF, Supplemental file 1**). The survey is completely anonymous: names or other personal identifiers (e.g. email-addresses, initials) will not be recorded and not included in any database. Participants are members of the general public, not from a patient population. The study is purely observational without any medical intervention, drug or medical device administration, and will be conducted in accordance with the ethical principles outlined in the Declaration of Helsinki.

This study was reviewed by Salus IRB and determined to be exempt from ethical approval on February 13, 2025. The exemption was granted in accordance with U.S. federal regulations under 45 CFR 46.104(d)(2)(i)(ii).

## Appendix 1 Adaptations of patient-reported outcomes to the general population

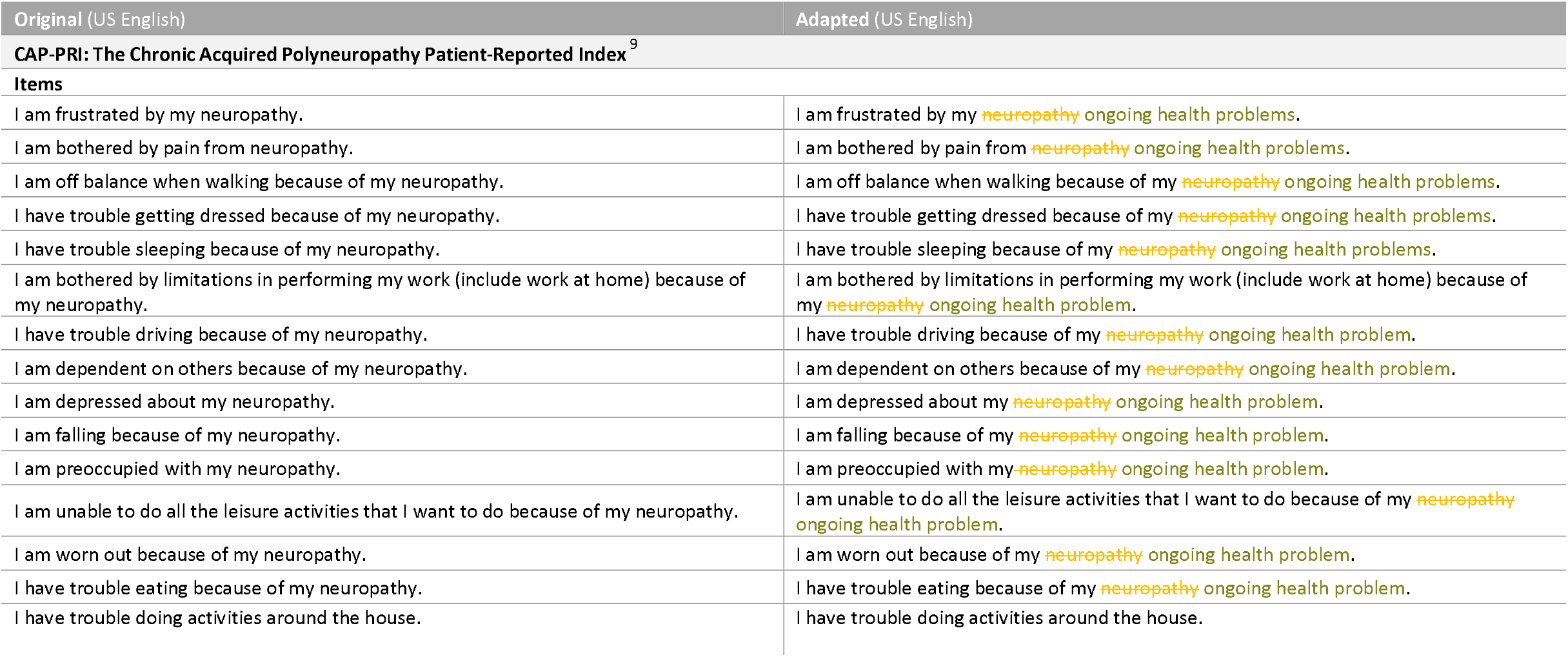
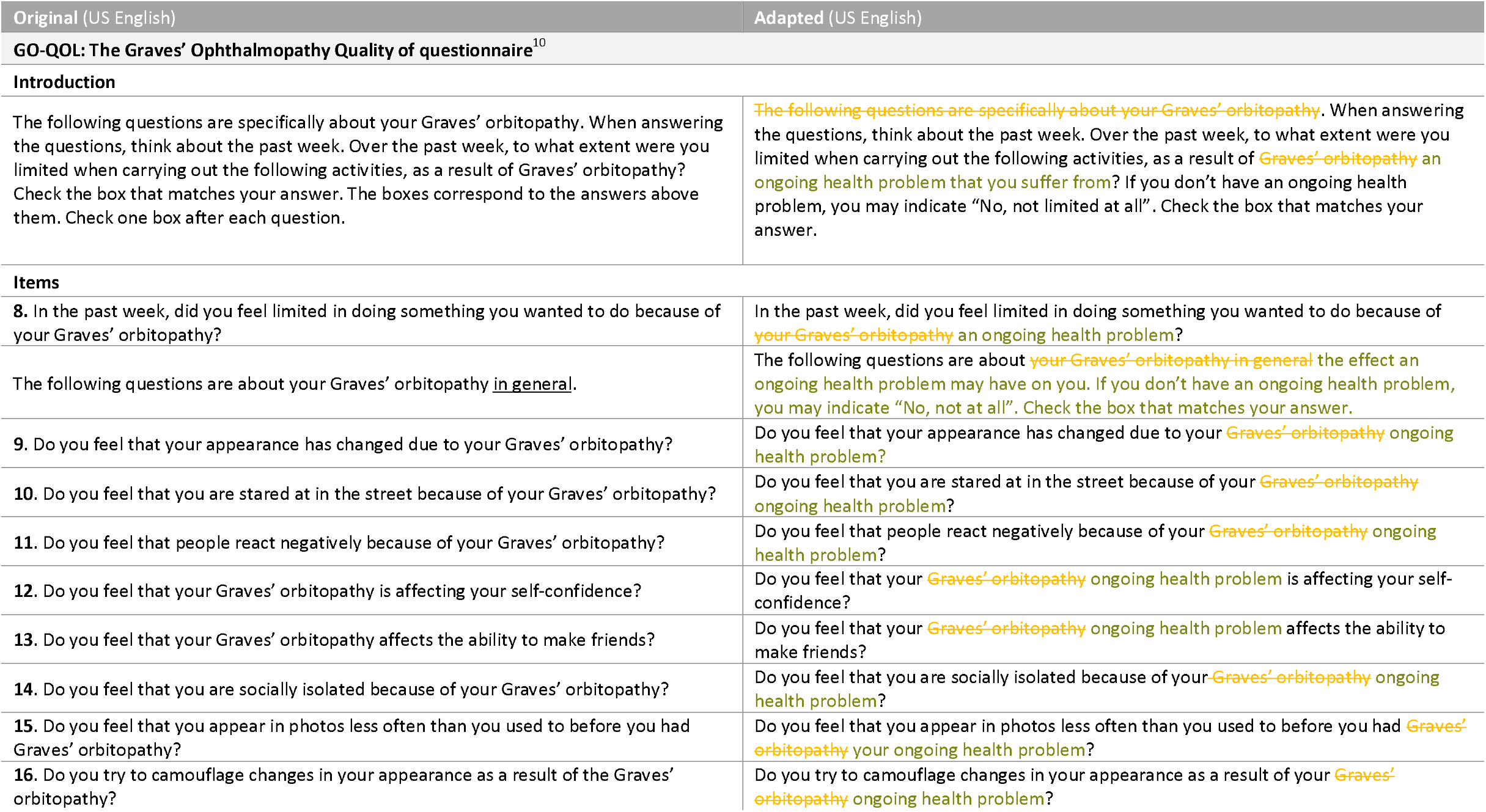

## Appendix 2 Experimental Arm: EQ-5D-5L Bolt-ons and Modified Composite Dimensions

In addition to the standard EQ-5D-5L questionnaire included in the GENESIS study, an experimental arm was added to investigate the psychometric properties of additional dimensions. This methodological research consists of two parts: (1) evaluation of five vitality-related EQ-5D-5L bolt-ons, and (2) comparison of composite versus component dimensions. All participants will complete the standard EQ-5D-5L, after which they will be randomized into two groups. Half of all participants will complete the bolt-ons, and the other half the modified composite dimensions.

### 1. Vitality-related EQ-5D-5L Bolt-ons

#### Background

The EQ-5D-5L is a widely used instrument for assessing health-related quality of life (HRQoL), but its five core dimensions may not fully capture dimensions relevant to both patients and the general population.

#### Objectives

To address this, five vitality-related bolt-ons will be evaluated: tiredness, energy, exhaustion, and sleep. The specific research questions are:

1. What are the comparative psychometric properties of these five bolt-ons in the general population?
2. How do they relate to each other and to validated fatigue measures (FACIT-Fatigue and RT-FSS)?
3. Which bolt-on is most preferred in terms of content relevance?
4. Does psychometric performance vary across countries and languages?

#### Methods

Directly after completing the standard EQ-5D-5L, half of all participants will be shown the five bolt-ons, followed by a ranking exercise. To ensure consistency, the levels, wording, and lay-out of the bolt-ons are very similar to the standard EQ-5D descriptive system. The order of bolt-ons will be randomized for every participant to reduce bias related to the ranking of the bolt-ons.

This analysis is based on data from wave 1 and wave 2 of the GENESIS study across six countries. Wave 1 includes all bolt-on items and fatigue-specific PROMs. In wave 2 (a 2-month follow-up), test-retest reliability (Gwet’s AC, weighted kappa) and responsiveness will be assessed. Psychometric evaluation includes distributional characteristics (floor/ceiling effects, profile counts), informativity (Shannon H’ and J’), divergent and convergent validity, explanatory power (linear regression predicting EQ VAS), and structural validity (CFA, item response theory). Measurement invariance across five languages (English, Spanish, German, Italian, Japanese) will also be tested.

### 2. Modified composite dimensions

#### Background

The EQ-5D-5L includes one dimension covering anxiety and depression, and one clustering pain and discomfort. Similarly, the EQ-5D-Y (Youth) version groups being sad, worried or unhappy in one dimension. However, these composite dimensions may mask distinctions between their components.

#### Objectives

The aim is to investigate the performance of individual health descriptors compared to composite dimensions. Specific research questions include:

1. How do the composite items compare to the components?
2. How do the six individual mental health items relate to each other?
3. What are the psychometric properties of these dimensions in the general population?
4. Do these properties differ by language group?

#### Methods

Immediately following the standard EQ-5D-5L, half of all participants will be presented with a block of modified composite dimensions. The composite “pain/discomfort” and “anxiety/depression” dimensions remain part of the standard EQ-5D-5L, while the combined “worried/sad/unhappy” item—originating from the EQ-5D-Y—was added to this block of modified composite dimensions. In this block, all composite dimensions have been carefully split up, isolating only one component in the dimension name and its levels. Finally, the item “happy” was added to see what the impact is of removing double negatives in the “unhappy” dimension (e.g. “I am not unhappy”).

Data will be drawn from wave 1 and wave 2 of the GENESIS study. We will examine response patterns, and apply psychometric techniques including factor analysis, as well as Item Response Theory (IRT) models, to assess whether individual and composite descriptors load on the same latent factor structure. Psychometric analyses include distributional characteristics, informativity (Shannon H’ and J’), convergent and divergent validity, explanatory power, test-retest reliability, and responsiveness. The Hospital Anxiety and Depression Scale (HADS) was included to assess convergent validity. Measurement invariance across languages will be assessed using standard confirmatory factor analysis models and thresholds for fit change.

