## Supplementary file 1 for "The Study Protocol for GENESIS: GENEral population normS—An International Survey"

**Informed Consent Form**

**Sponsor:**	argenx BV

**Principal Investigator:**	Services in Health Economics

Boulevard Lambermont 418

1030 Schaarbeek

Brussels-Capital Region

Belgium

**Contact information:**	[**Contact form**](https://www.she-consulting.be/contact-us/)

### Research Consent Summary

You are invited to take part in an online survey about your health, daily life, and use of medical services. Your answers will be anonymized, stored securely, and used only for research, in compliance with data protection and cybersecurity regulations (GDPR and NIS-2). Participation is voluntary, and you may stop at any time. As a thank-you, you will receive points that can be redeemed for gifts.

**If you have questions, concerns, or complaints talk to the research team via the contact information listed above.**

**Detailed Research Consent**

### Why is this research being done?

We want to compare data on health, functioning in daily life, work, social life, and use of medical services between the general population and people with rare diseases. About 21,000 respondents from Germany, Italy, Japan, Spain, the UK and US will take part in this survey.

### What happens if I agree to take part in this research?

You will be directed to an online survey that includes questions about your demographic profile, health-related quality of life, and asks about your physical functioning, your mental well-being, any pain or fatigue you may be experiencing, your possible need for help from a caregiver, and whether you had to take time off work due to illness. Participation will last no longer than the time it takes to complete the survey (on average, 12 minutes).

### Could being in this research hurt me?

No, the survey is anonymous and involves only answering questions.

### Will being in this research benefit me?

No, but this research aims to provide valuable data for patients suffering from rare diseases.

### What happens to the information collected for this research?

Your responses will be completely anonymized, and your personal information will not be collected. Your data will be combined with data from other respondents. The data will be securely stored and used solely for research purposes. The study complies with GDPR and the NIS-2 Directive in terms of personal data protection and cyber security.

### Who can answer my questions about this research?

If you have questions, concerns, or complaints, talk to the research team at the phone number listed in this document, or write an email. It is also possible to leave a comment in the free text box at the end of the survey.

This research is being overseen by Salus IRB. An IRB is a group of people who perform independent review of research studies. You may contact them at +1 512-380-1244 or.com if you cannot reach the research team.

### What happens if I agree to be in this research, but I change my mind later?

Your participation in this study is voluntary. You may decide not to participate, and you have the possibility to leave the study at any time.

**Will I be paid for taking part in this research?**

Yes, you will receive an incentive in the form of points that can be exchanged for gifts through the online panel that recruited you.

**Agreement to Participate in the Survey**

I understand all the information provided here and my de-identified data may be used in any type of quality of life and healthcare research. I am 18 years or older and I participate on a voluntary basis.

- I agree to participate
- I do not agree to participate

Thank you for your time!

OR

We're sorry that you can't complete the survey. You can now close this window.
